# Identifying Predictors of Benzodiazepine Discontinuation in Medical Cannabis Patients with Post-traumatic Stress Disorder Using a Machine Learning Approach

**DOI:** 10.1101/2025.03.05.25323416

**Authors:** Mitchell L. Doucette, Mark Kasabuski, Emily Fisher, Junella Chin, Douglas Bruce, Panagiota Kitsantas

## Abstract

**Introduction:** Post-Traumatic Stress Disorder (PTSD) is a debilitating mental health condition commonly treated with medications like benzodiazepines (BZDs), despite their potential for negative long-term side effects. Medical cannabis has emerged as a possible adjunctive therapy for PTSD. However, the relationship between medical cannabis use, relief from PTSD symptoms, and the use of BZDs remains unclear. Thus, we sought to identify predictors of changes to BZD usage among medical cannabis patients with PTSD.

**Methods:** This study utilized survey data from PTSD patients in the Leafwell patient database, collected in the fall of 2023. To assess the relationship between medical cannabis use, PTSD symptom relief, and the discontinuation of BZDs, we employed a multi-step analysis approach. First, we developed a decision tree model to identify key predictors of BZD discontinuation, including prior cannabis use and reported PTSD relief post-medical cannabis treatment initiation. The tree was pruned using the optimal complexity parameter to improve model interpretability. Following this, a secondary logistic regression analysis was performed to confirm the significance of key predictors identified by the decision tree.

**Results:** In the pruned decision tree, not currently receiving psychiatric care for their PTSD was the strongest predictor of BZD discontinuation, followed by self-reported efficacy of medical cannabis in relieving PTSD symptoms, prior cannabis use, and history of traumatic brain injury (TBI) among medical cannabis patients. Age was also a significant factor, with younger individuals more likely to discontinue. Logistic regression analysis supported these findings, with receiving care, TBI, and cannabis use remaining key predictors. Interaction models suggest prior cannabis use moderates the relationship between those not receiving psychiatric care and BZD discontinuation odds.

**Discussion:** These findings suggest that medical cannabis may offer a promising route for BZD discontinuation for long-term users with PTSD symptoms. The association between cannabis use and BZD discontinuation highlights the need for further exploration of cannabis as an adjunctive therapy in PTSD care. More research is necessary to confirm the long-term safety and effectiveness of medical cannabis in this context, ensuring that it can be integrated into care without unintended negative consequences. Individualized care approaches remain crucial given varying patient factors.

## 1 Introduction

In 2020, research estimated that approximately 13 million Americans were living with a post-traumatic stress disorder (PTSD) (National Center for PTSD, 2023). It is estimated about 6 out of 100 Americans are diagnosed with PTSD during their lifetime, with women (8 out of every 100 women) more likely to be diagnosed than men (4 out of every 100 men) (National Center for PTSD, 2023). This potentially incapacitating mental health disease can arise from either experiencing or witnessing a traumatic event (Bryant, 2019). Symptoms of PTSD vary from person to person and can include disturbed sleep and nightmares, flashbacks to the traumatic event, difficulty concentrating, etc. In the United States (US), veterans have a heightened risk of PTSD compared to the general population (Christiansen and Berke, 2020).

### 1.1 Benzodiazepines as a Treatment for Post-traumatic Stress Disorder

PTSD is often treated through a combination of psychotherapeutic, counseling, and pharmacologic interventions aimed at decreasing symptoms and improving functional outcomes. Pharmacological treatments, particularly selective serotonin reuptake inhibitors (SSRIs), have demonstrated effectiveness in managing PTSD symptoms (Williams et al., 2022) as have benzodiazepines for short-term relief. Benzodiazepines (BZD) are a class of medication approved to treat generalized anxiety disorder, insomnia, seizures, social phobia, and panic disorder.

BZD medications have become one of the most commonly prescribed drug classes in the United States (US), with an estimated 92 million prescriptions dispensed in 2019 (U.S. Food & Drug Administration, 2024). Between 30-74% of patients with PTSD are prescribed a BZD (Guina et al., 2015). Their medicinal properties can be advantageous for immediate symptom control; however, the long-term use of benzodiazepines is associated with several significant drawbacks. Side effects include the impairment of cognitive and motor functions, risk of developing tolerance, physical dependence, and addiction. After opioids, BZD are the medication most involved in prescription overdose deaths (National Institute on Drug Abuse, 2024; Maust et al., 2023). In 2018, an estimated 50% of patients who were dispensed oral BZD received them for a duration of two months or longer (Substance Abuse and Mental Health Services Administration, 2019).

BZD are primarily prescribed for short-term use due to their potential for dependence, withdrawal symptoms, and long-term side effects. These medications, while effective for acute anxiety and insomnia, pose significant risks when used over extended periods, including cognitive impairment, tolerance, and addiction (Reid Finlayson et al., 2022). The Veterans Affairs guidelines highlight that prolonged BZD use can interfere with the effectiveness of trauma-focused therapies like cognitive-behavioral therapy for PTSD, further discouraging long-term use (U.S. Department of Veterans Affairs, 2022). Additionally, Allary and colleagues (2020) note that patients may struggle with discontinuation after long-term use, experiencing severe withdrawal symptoms that can mimic or exacerbate the original condition (Allary et al., 2020).

Moreover, discontinuing BZD treatment poses a significant challenge for a substantial proportion of patients (Maust et al., 2023; Reid Finlayson et al., 2022; Takamine et al., 2024; Takeshima et al., 2022; Fernandes et al., 2022). Reid Finlayson et al. (2022) report that 63.2% of individuals who had taken BZD struggled with discontinuation, while Maust et al. (2023) indicate that nearly half of long-term users fail to successfully taper off these medications. The difficulty largely stems from the development of physiological dependence, which can lead to withdrawal symptoms such as anxiety, insomnia, and cognitive impairments. These symptoms often mimic the original reasons for which the medication was prescribed, creating a cyclical challenge that makes tapering off particularly difficult. Furthermore, other research among older adults suggests that psychological factors, such as fear of withdrawal symptoms and lack of confidence in the tapering process, exacerbate these difficulties (Allary et al., 2020). The unpredictable nature of symptom recurrence, compounds these challenges, making discontinuation a protracted and complex process for many patients.

The long-term risks and dissatisfaction with BZD treatments may lead PTSD patients to seek alternative and complementary therapy. Some of these therapies include yoga, mindfulness exercises, acupuncture, Tai Chi, and herbal medications have varying degrees of efficacy and validity (Rehman et al., 2021; Wynn, 2015; Boyd et al., 2018; Numata et al., 2014; Tangkiatkumjai et al., 2020). An emerging treatment option for PTSD is medical cannabis.

### 1.2 Medical Cannabis and Post-traumatic Stress Disorder

PTSD qualifies for medical cannabis use in 36 out of 38 US states and is frequently reported by patients as a qualifying condition in states where cannabis is solely for medical purposes (Boehnke et al., 2019). In the body, cannabis exerts its pharmacological effects via interaction with the endocannabinoid system (ECS) (Hill and Gorzalka, 2009). Given the complexity of the ECS, this discussion will focus on elements that research has linked to the clinical pathophysiology of PTSD, particularly the CB1 receptor and its two endogenous ligands: N-arachidonoylethanolamide (AEA, also known as anandamide) and 2-arachidonoylglycerol (2-AG).

The hypothalamic-pituitary-adrenal (HPA) axis and the sympathetic nervous system could represent a key connection between the development of PTSD and potential ECS involvement. Current findings suggest that individuals exposed to trauma, either immediately or shortly thereafter, show reduced cortisol levels, likely due to increased glucocorticoid receptor sensitivity (Pervanidou and Chrousos, 2010). This cortisol reduction leads to heightened arousal from increased noradrenergic transmission, which may contribute to PTSD onset. Research by Yehuda (2009) and Sarapas et al. (2011) pointed to glucocorticoid signaling as a potential genetic marker for PTSD. The ECS appears responsive to glucocorticoid hormones, which may help regulate aspects of the stress response, specifically through the feedback mechanism that terminates HPA axis activity (Di et al., 2003; Hill et al., 2010; Evanson et al., 2010; Hill et al., 2011).

Research using a population-based cohort near the 9/11 events, revealed that PTSD is linked to lower circulating levels of 2-AG, and both endogenous CB1 receptor ligands (2-AG and AEA) were associated with specific PTSD symptom clusters, particularly the retention of negative emotional memories (Hill et al., 2013). This suggests that cannabinoid-based therapies could be effective in managing certain PTSD symptoms.

There is an increasing amount of literature that supports the use of medical cannabis for managing PTSD, although much of the evidence comes from studies that do not utilize randomized control trials (Rehman et al., 2021; Pillai et al., 2022; Lynskey et al., 2024; Cahill et al., 2021; Sznitman et al., 2022; Nacasch et al., 2023; Roitman et al., 2014; Krediet et al., 2020; LaFrance et al., 2020). Various studies have reported reductions in PTSD symptom severity and improvements in sleep quality after initiating medical cannabis treatment, with only minimal adverse effects observed in patients (Pillai et al., 2022; Lynskey et al., 2024; Cahill et al., 2021; Sznitman et al., 2022; Nacasch et al., 2023; Krediet et al., 2020). Additionally, some research has focused on how medical cannabis affects the overall quality of life for individuals, with promising outcomes in enhancing well-being (Pillai et al., 2022; Cahill et al., 2021). While only a few studies have specifically targeted PTSD patients in this regard, the positive effects on quality of life are notable. These findings suggest that medical cannabis may play a beneficial role in managing PTSD-related symptoms and improving patients’ daily functioning.

### 1.3 Current Contribution

Limited research exists on the use of medical cannabis and BZD discontinuation within the context of PTSD. The study by Purcell et al. (2019) found that among 146 patients prescribed medical cannabis, 45.2% successfully discontinued BZD use after an average of six months. Discontinuation rates were highest after the first follow-up (30.1%) and continued to increase by the second visit (44.5%), but showed minimal change thereafter. The study suggests a significant association between medical cannabis therapy and reductions in BZD use, warranting further exploration of cannabis as an alternative treatment option. Two other studies have examined the use of medical cannabis and BZD usage; however both were within the context of chronic pain management (O’Connell et al., 2019; Kalaba et al., 2022).

To date, no research has investigated what demographics or clinical characteristics may drive BZD discontinuation within the medical cannabis patient population with PTSD. To this end, we sought to understand: 1) Determine the proportion of PTSD patients using medical cannabis who were prescribed a BZD, and among those, assess how many reduced or discontinued their BZD use after initiating medical cannabis treatment; 2) Identify demographic and clinical characteristics that predict BZD discontinuation; and, 3) Examine whether prior cannabis use moderates the relationship between patient characteristics and the likelihood of BZD discontinuation.

## 2 Methods

### 2.1 Data

We conducted a cross-sectional survey of medical cannabis patients using the Leafwell Patient Database (Doucette et al., 2024). Leafwell, Inc. is a telemedicine portal that facilitates the acquisition of medical cannabis cards across 36 states in the U.S. Leafwell randomly selected 50% of its patients who received authorization to obtain a medical card because of their PTSD from January 1, 2020 to October 1, 2023 (n = 9,168) to complete an online anonymous survey that included demographic indicators, use of medical cannabis, use of prescription medication and psychiatric and psychological care, and PTSD symptomology. Recruitment involved an initial recruitment email sent in early November of 2023 with a follow up email sent out 2 weeks later. Participants who completed the survey were invited to enter a raffle for thirty $50 Amazon gift cards. The survey took 5-7 minutes to complete, and a total of 970 participants completed the survey (response rate = 10.6%). The survey was executed using Qualtrics. The study protocol was reviewed and approved by the Institutional Review Board at DePaul University.

In an effort to assess non-response bias, we compared limited demographic characteristics of respondents and non-respondents (see Supplemental Table 1) using available information from the Leafwell Patient Database. Among respondents (n = 970), the mean age was 40.02 years (SE = 0.44), which did not differ significantly from non-respondents (n = 8,198) (39.32 years, SE = 0.10; p = 0.9323). Veteran status also did not differ (p = 0.676). However, a higher proportion of respondents were male (65.1% vs. 44.9% among non-respondents; p < 0.001), and respondents were more likely to be White, non-Hispanic (73.8% vs. 68.4%; p = 0.001). Collectively, these findings suggest that our respondent pool may over represent male and White, non-Hispanic PTSD patients compared to the broader group of potential participants.

### 2.2 Survey Questions

To understand BZD usage, participants were asked a series of yes-or-no questions. First, participants were asked, “Prior to starting medical cannabis, were you prescribed or taking benzodiazepines (e.g., lorazepam, clonazepam, alprazolam) or other anti-anxiety medications?” If they responded “Yes” to the question about BZD prescription, they were then asked two follow-up questions: “Has your use of medical cannabis allowed you to reduce your use of benzodiazepines or other anti-anxiety medications?” and “Has your use of medical cannabis allowed you to discontinue or stop your use of benzodiazepines or other anti-anxiety medications?”

We collected demographic measures including gender (Male vs. Female), age, and race/ethnicity (Black, non-Hispanic, White, non-Hispanic, Hispanic, Other racial/ethnic identity). Other measures included education level (Less than high school, High School/GED, Bachelor’s degree, or Master’s degree or above), military veteran status (Yes/No), and state of residence. We collapsed the state of residence into a binary variable that captured whether the patient lived in a state with medical cannabis only or medical cannabis and recreational cannabis.

Participants were asked to complete three brief clinical measurements on health-related topics. First, patients were asked about the severity of their PTSD using the Impact of Event Scale-6 scale (IES-6), a commonly used measures of posttraumatic stress reactions (Thoresen et al., 2010). Using established guidelines, we categorized patients into clinical PTSD (defined as IES-6 score >=7) and sub-clinical PTSD (defined as IES-6 score <=6) based on previous research (Thoresen et al., 2010). Patients were also asked about the severity of their anxiety (Generalized Anxiety Disorder (GAD) scale-7) and insomnia (Insomnia Severity Index (ISI)). For GAD-7, anxiety severity was categorized as minimal anxiety, mild anxiety, moderate anxiety, and severe anxiety per GAD-7 guidelines (Spitzer et al., 2006). For ISI, insomnia severity was categorized into no insomnia, subclinical insomnia, moderate insomnia, and severe insomnia per ISI guidelines (Morin et al., 2011). Participants were also asked two cannabis utilization measures, including history of cannabis use prior to obtaining medical cannabis treatment (Question = “Did you use marijuana or cannabis before you received a medical marijuana card or medical marijuana registration?”) and past week use of cannabis (Question = “Think specifically about the past 7 days up to and including today. On how many days did you use cannabis?”). We categorized past week use of cannabis into a binary variable representing whether a patient used medical cannabis every day (Yes) or less than every day within the past week (No).

In addition, participants were asked about past medical history relevant to their PTSD, including history of traumatic brain injury (Yes/No), experience of interpersonal violence (Yes/No), and whether they were currently receiving psychiatric care (Yes/No), individual therapy (Yes/No), or current group therapy (Yes/No). Participants also reported the length of their PTSD symptoms (0-6 months, 7-12 months, 1-2 years, 2-5 years, more than 5 years). We also asked participants to self-report the extent to which medical cannabis had relieved their PTSD symptoms, categorized into one of the following: not relieved, somewhat relieved, moderately relieved, completely relieved. Due to a low number of participants responding ‘not relieved’, we collapsed ‘not relieved’ and ‘somewhat relieved’, creating a three-level categorical variable.

### 2.3 Analytic Approach

To answer our first research question, descriptive statistics were used to determine the percent of patients who reported discontinuing their use of BZD medications. Associations between demographics, clinical scales, medical history, cannabis use/history, and discontinuation of BZD medications were assessed using the chi-squared test. The t-test was used to examine any significant differences in the average age between those who discontinued BZD compared to those who did not.

To answer our second research question about demographic or clinical characteristics that may predict BZD continuation, we conducted a two-step decision tree analysis. Decision tree analysis is a non-parametric method for classification. It allows for the identification of key predictors through recursive partitioning of the data. By splitting the data into meaningful subsets based on the most influential variables, decision trees provide an interpretable framework for understanding how various factors contribute to an outcome. This method is particularly useful in clinical settings, as it enables the visualization of decision-making pathways and, relevant to the scope of this study in identifying significant clinical factors that influence BZD discontinuation.

Our initial decision tree contained all of the variables considered in this study (see Table 1). Once constructed, we performed cross-validation to identify the complexity parameter (CP) that minimized the cross-validation error, which helps the model with overfitting. Pruning the decision tree is best practice in decision tree analysis as it reduces model complexity and improves interpretability and generalizability by removing non-informative splits. As this is a relatively novel area of research, we selected a CP that provides a modest level of tradeoff between model complexity and prediction accuracy.

**Table 1:**
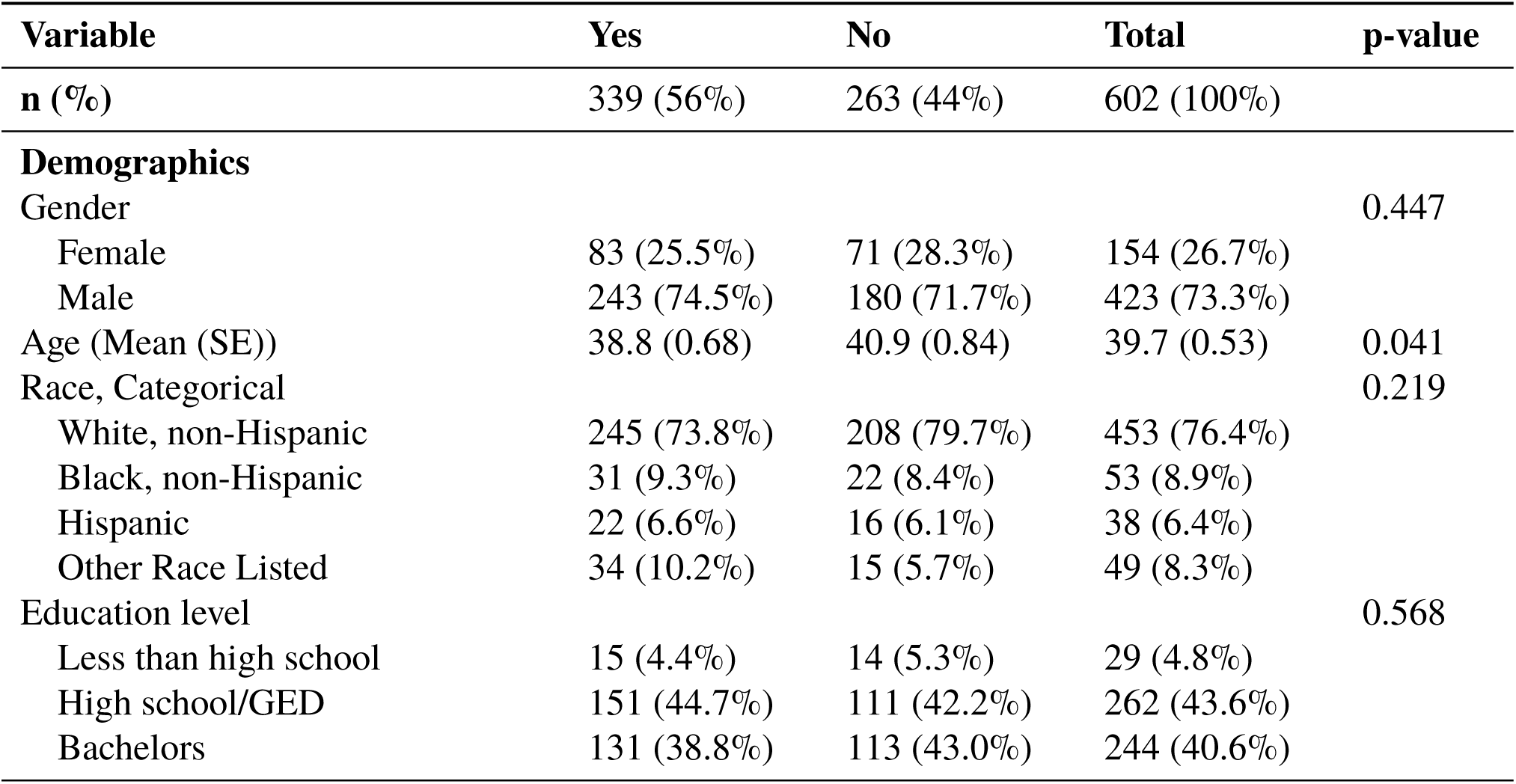
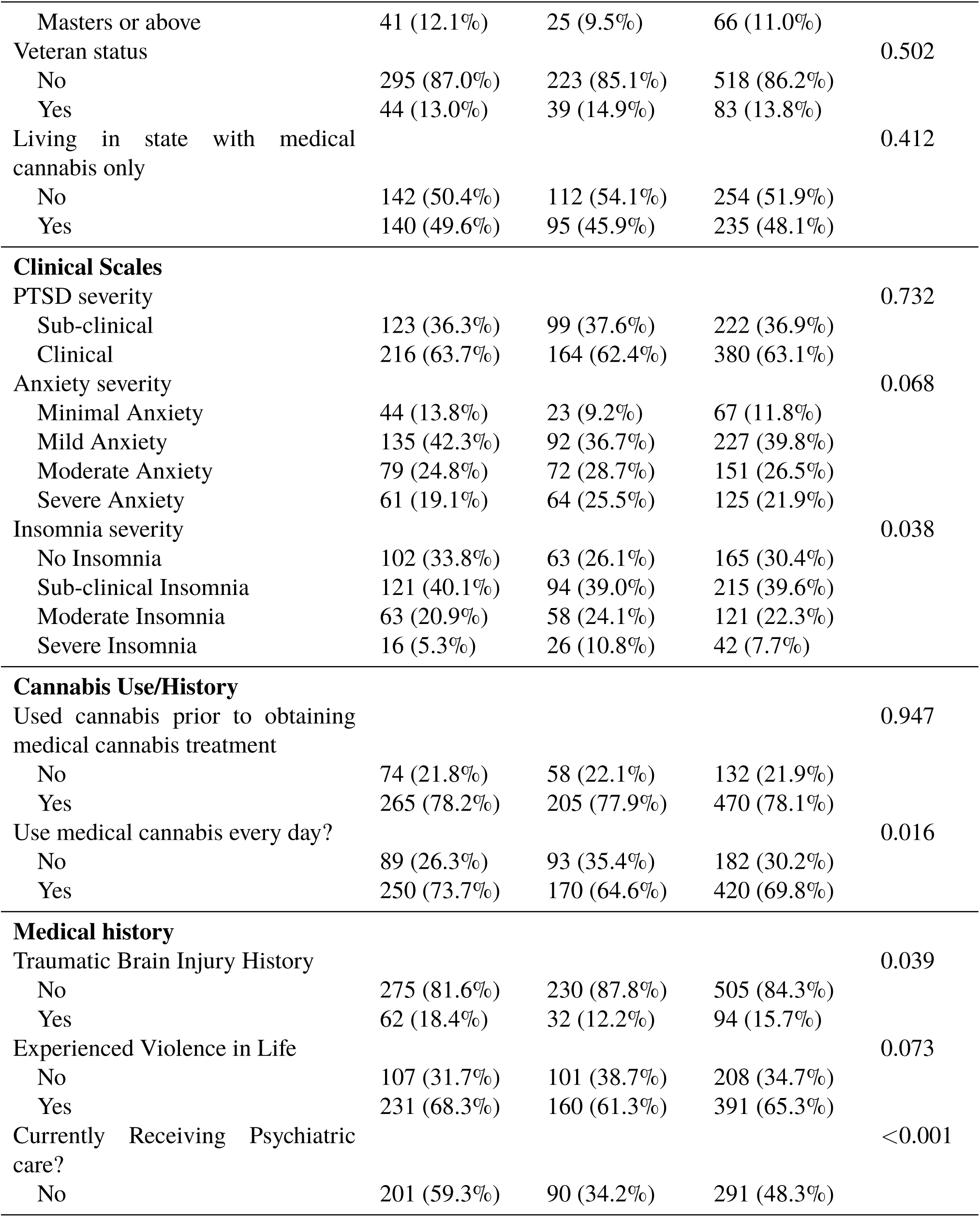
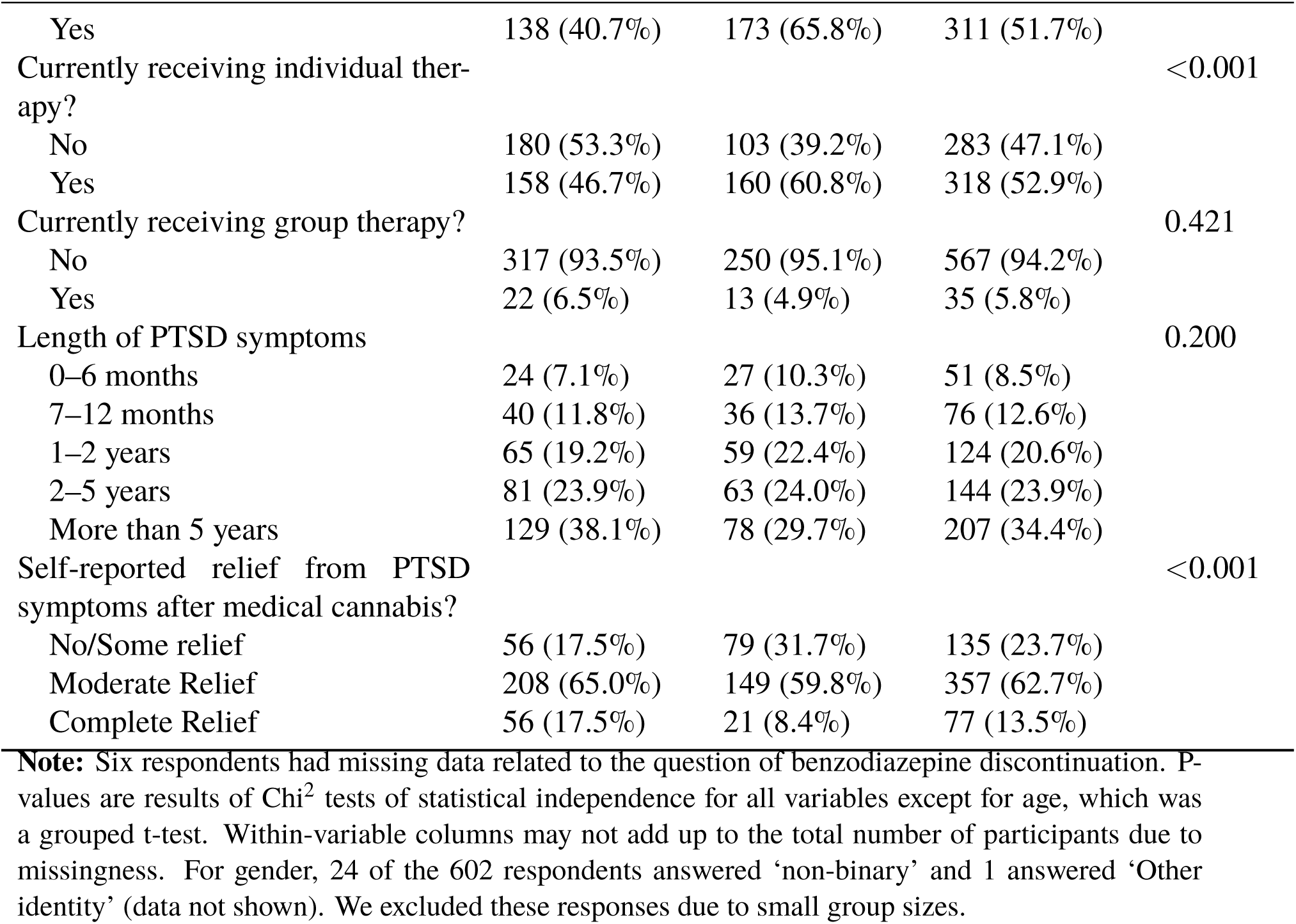
Associations between sample characteristics and benzodiazepine discontinuation status after starting medical cannabis treatment among patients with post-traumatic stress disorder.

To answer our third research question (whether prior cannabis moderates the relationship between characteristics and the likelihood of discontinuing BZD), we conducted a logistic regression using the predictors found to have a significant impact on the probability of BZD medication discontinuation within the pruned decision tree. Within the logistic regression, we included an interaction term between our binary variable of prior cannabis use and the most important predictor of BZD discontinuation. We evaluated the predicted probabilities of the categorical variables to understand if cannabis use prior to starting medical cannabis treatment moderated the relationship between the most important predictor of BZD discontinuation. The logistic regression model included robust standard errors.

As part of model diagnostics of the decision tree analysis, we conducted a permutation test to assess the importance of the individual predictors included in the initial decision tree. The permutation randomly shuffles the values of a particular variable in the dataset while keeping all other variables unchanged. The decision tree is then applied to this modified dataset, and the model’s accuracy is recalculated. By comparing the accuracy of the model on the permuted data to the accuracy of the original data, the permutation test quantifies how much a variable contributes to the model’s predictive performance. Further, we provided model performance metrics for the pruned decision tree. Metrics include accuracy, no information rate, sensitivity and specificity, and positivity and negative predicted values as well as other tests of model balance and accuracy.

## 3 Results

Of the 970 total participants surveyed, a total of 608 patients reported using a BZD in the past six months (62.7%). Of the 608 patients that reported taking a BZD in the past six months, 339 (55.8%) reported discontinuing their treatment while 263 (43.3%) did not; Six additional participants did not respond (0.99%). Of the 263 participants that did not discontinue their BZD medication after starting medical cannabis, 75.7% (n = 199) reported reducing their use of the the medication (data not shown). Otherwise said, 538 of the 608 patients that reported taking a BZD (88.4%) either discontinued or reduced their use of BZD.

Table 1 provides cross-tabulations examining demographic and clinically relevant characteristics across patients who discontinued their BZD vs. patients who did not. Gender was not significantly related with BZD discontinuation (p = 0.447), although the majority of participants were male (73.3%). Education (p = 0.568), veteran status (p = 0.502), and race/ethnicity (p = 0.219) were also not significantly related to BZD discontinuation. Age, however, was significantly different between the groups, with those discontinuing BZD use being slightly younger (mean age 38.8 years vs. 40.9 years; p = 0.041).

Characteristics revealed that prior cannabis use did not influence BZD discontinuation (p = 0.947), though a higher percentage of daily cannabis users reported discontinuing compared to non-daily users (73.7% vs. 64.6%; p = 0.016). PTSD severity (p = 0.732) and anxiety severity (p = 0.068) were not statistically different, although patients with minimal anxiety trended towards discontinuation. Length of PTSD symptoms likewise did not display a statistically significant association with BZD discontinuation. However, a history of traumatic brain injury (TBI) (p = 0.039) was significantly associated with higher rates of discontinuation.

For psychiatric care, a higher percent of patients who received some type of care related to their PTSD discontinued their BZD (65.8% vs. 40.7%; p < 0.001). Engagement in individual therapy was positively associated with discontinuation (p < 0.001), while group therapy showed no significant association (p = 0.421). Notably, patients who perceived medical cannabis as effective for relieving PTSD symptoms were more likely to discontinue BZDs (p < 0.001).

### 3.1 Decision Tree Results

The initial decision tree model (See Supplemental Figure 1), with a root node error of 0.442, was subjected to cross-validation to identify an optimal level of pruning. The cross-validation plot (Supplemental Figure 2) indicated a point where the relative error stabilized, suggesting the model was no longer benefiting from additional splits. By setting the CP to 0.015210, we reduced the model’s complexity from 11 splits in the unpruned tree to a more parsimonious 5-split model while keeping the error rate around 0.75 (relative error = 0.776).

The pruned decision tree (See Figure 1) identified several key variables that influence the likelihood of BZD discontinuation among medical cannabis patients. The primary split was based on whether the patient was receiving psychiatric care. Nearly half (48%) of patients who did not receive psychiatric care discontinued their BZD medications, suggesting that those engaged in psychiatric care may be more likely to continue use

**Figure 1:**
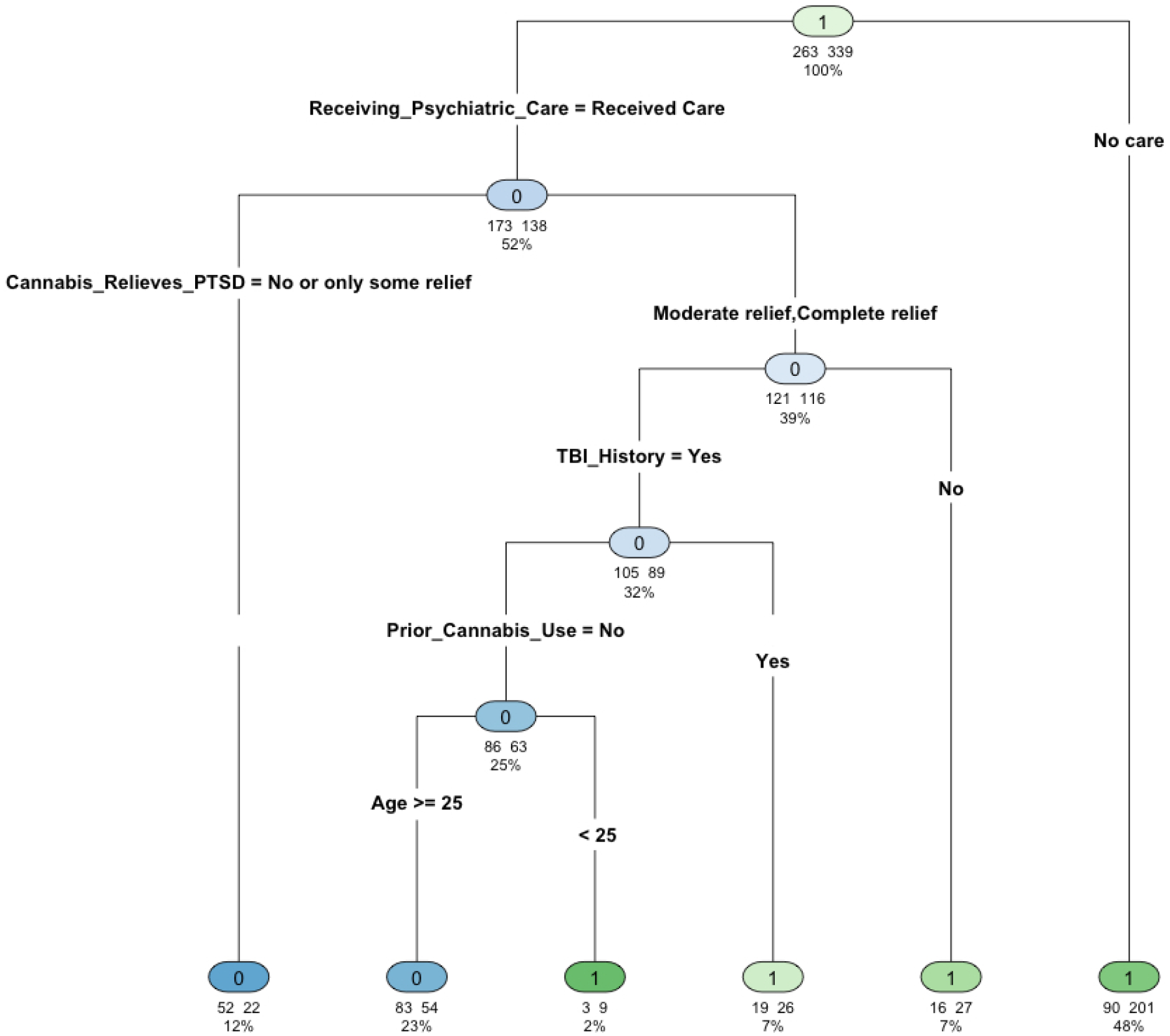
Pruned Tree decision tree estimating the most influential predictors of benzodiazepine discontinuation among medical cannabis patients with post-traumatic stress disorder. Each circle (node) in the tree represents a decision point based on one of the predictor variables. The text or label above/below the circle indicates the variable and the specific threshold or category used to split the sample. The first line of numbers beneath each circle shows how many participants fall into each branch at that decision point. The percentage shown under those numbers indicates the proportion of the overall sample (or that node’s sub-sample) captured at that branch.

Among patients who received psychiatric care, the next significant factor was their perception of medical cannabis’s effectiveness in relieving PTSD symptoms. Those who reported no or only some relief had a lower discontinuation rate (12%), indicating that perceived inefficacy of cannabis may be associated with continued BZD use. In contrast, patients who experienced moderate or complete relief from cannabis were more likely to discontinue, though additional factors further influenced their likelihood of stopping BZD treatment.

For those who reported moderate or complete relief, traumatic brain injury (TBI) history emerged as an important predictor. Patients without a history of TBI had a higher likelihood of discontinuing, whereas among those with a TBI history, prior cannabis use became a distinguishing factor. Patients with no prior cannabis use were further stratified by age, with younger patients (under 25) more likely to discontinue compared to those 25 and older. Among patients with both a history of TBI and prior cannabis use, discontinuation rates remained high regardless of age.

### 3.2 Logistic Regression Results

The logistic regression analysis examined factors associated with BZD discontinuation among medical cannabis patients and is summarized in Table 2. Our final model included the characteristics that were found to significantly predict BZD discontinuation in the pruned decision tree. The model included TBI history, prior cannabis use, receipt of psychiatric care for their PTSD, perceived efficacy of medical cannabis for PTSD, age, and an interaction term between prior cannabis use and psychiatric care.

**Table 2:** Logistic regression model examining significant predictors of benzodiazepine discontinuation among medical cannabis patients with post-traumatic stress disorder.

| Variable | Odds Ratio | 95% CI | p-value |
| --- | --- | --- | --- |
| Traumatic Brain Injury History | 2.25 | 1.28, 3.97 | 0.005*** |
| Used cannabis prior to obtaining medical cannabis treatment | 1.67 | 0.84, 3.33 | 0.146 |
| Currently Receiving Psychiatric Care? | 0.93 | 0.40, 2.19 | 0.869 |
| **Self-reported relief from PTSD symptoms after medical cannabis:** |  |  |  |
| Moderate Relief vs. No/Some Relief | 1.99 | 1.21, 3.27 | 0.007** |
| Complete Relief vs. No/Some Relief | 4.19 | 2.00, 8.77 | <0.001*** |
| Age | 0.98 | 0.96, 0.99 | 0.013* |
| Interaction term | 0.34 | 0.13, 0.89 | 0.029* |
| Intercept | 1.72 | 0.61, 4.81 | 0.304 |

**Note:** Significant codes for p-values: 0.001 \*\*\* 0.01 \*\* 0.05 \* 0.1 . 1. Model was a generalized linear model with binary and robust standard errors. Predictors included based on the final pruned decision tree (see Figure 1).

A significant association was observed between TBI history and BZD discontinuation. Patients with a history of TBI had over twice the odds of discontinuing BZDs compared to those without a TBI history (OR = 2.25, 95% CI: 1.28–3.97, p = 0.005). The perceived efficacy of medical cannabis for PTSD was also significantly associated with discontinuation. Compared to patients who stated their medical cannabis either did not relieve or only somewhat relieved their PTSD symptoms, patients who rated cannabis as moderately relieving their PTSD symptoms had nearly twice the odds of discontinuing (OR = 1.99, 95% CI: 1.29–3.27, p = 0.007), while those rating it as completely relieving were more than four times as likely to discontinue BZDs (OR = 4.19, 95% CI: 2.00-8.77, p < 0.001). Age was inversely associated with discontinuation, where each additional year was associated with a slight reduction in the odds of discontinuing (OR = 0.98, 95% CI: 0.96–0.99, p = 0.013).

Notably, the interaction between prior cannabis use and receiving psychiatric care was significant (OR = 0.34, 95% CI: 0.13–0.89, p = 0.029) (Table 2) This suggests that for patients who had used cannabis prior to obtaining a medical card and were also receiving psychiatric care, the odds of discontinuing BZDs were 66% lower compared to those without prior cannabis use and not receiving psychiatric care. In contrast, prior cannabis use on its own did not significantly predict discontinuation (OR = 1.67, 95% CI: 0.84-3.33, p = 0.148), nor did receiving psychiatric care (OR = 0.93, 95% CI: 0.40–2.19, p = 0.869), when considered independently, suggesting using cannabis prior to obtaining a medical card moderated the found relationship between those who received care for their PTSD.

Figure 2 provides the predicted probability of BDZ discontinuation for each category of the interaction. Among patients who did not use cannabis prior to starting medical cannabis treatment, there was no difference in the predicted probability of BZD discontinuation based on whether they had received psychiatric care (non-prior user, received psychiatric care, predicted probability (PP) = 0.529, 95% CI, 0.365–0.693; non-prior user, did not receive psychiatric care, PP = 0.580, 95% CI: 0.423–0.737). Prior cannabis use, before starting medical cannabis treatment, did moderate the relationship between receipt of psychiatric care and BZD discontinuation. Among prior cannabis users, those who did not receive care had a statistically significant higher PP (0.710, 95% CI: 0.610–0.810) compared to those who did receive care (0.441, 95% CI: 0.328–0.555).

**Figure 2:**
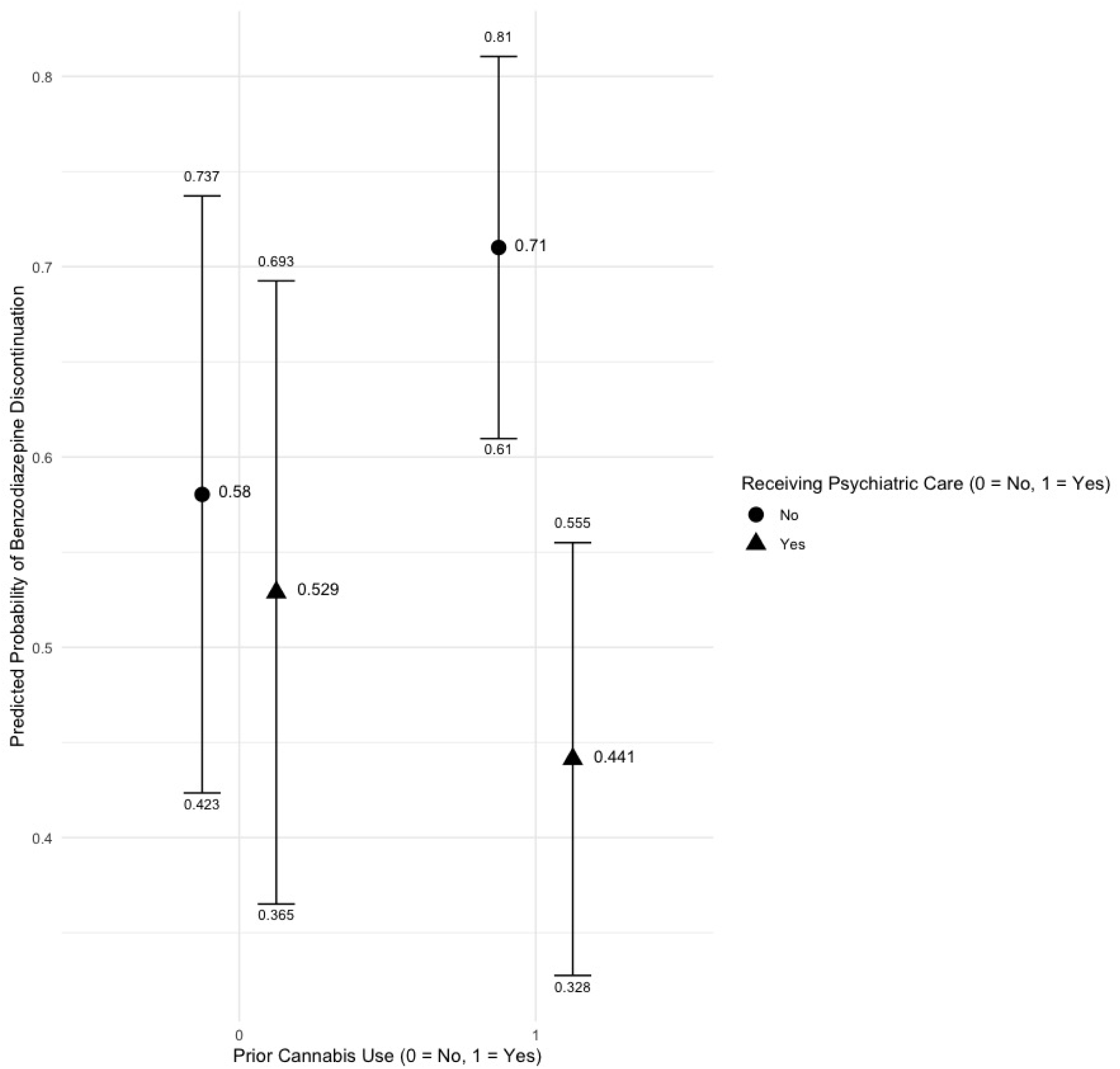
Predicted probabilities examining the interaction term between those who used cannabis prior to obtaining a medical cannabis card and those who were currently receiving psychiatric care.

### 3.3 Model Diagnostics

Supplemental Table 2 provides model performance metrics for the pruned decision tree. The pruned decision tree model demonstrated moderate predictive accuracy in identifying BZD discontinuation among medical cannabis patients. The overall accuracy of the model was 66.1% (95% CI: 62.2%, 69.9%), which significantly outperformed the no-information rate of 56.3% (p < 0.001). The Kappa statistic of 0.29 indicated moderate agreement beyond chance between the predicted and actual outcomes. Sensitivity and specificity were 51.3% and 77.6%, respectively, reflecting the model’s balanced ability to correctly identify both patients who did not discontinue (true negatives) and those who did discontinue (true positives). Positive and negative predictive values were similarly balanced, with the model correctly predicting non-discontinuation 63.9% of the time and discontinuation 67.3% of the time. McNemar’s test (p < 0.001) suggested that the misclassifications were equally distributed between the two classes. Overall, the balanced accuracy of 64.5% further supports the model’s moderately reliable performance in distinguishing between patients who discontinued and those who did not.

The permutation test results provided a detailed evaluation of variable importance in the decision tree model predicting BZD discontinuation among medical cannabis patients (See Supplemental Table 3) and heavily suggest our pruned decision tree is robust. The most influential variable in the model was receipt of psychiatric care with a mean accuracy reduction of 0.116 (95% CI: 0.080–0.148). Other variables that showed notable but smaller impacts on model accuracy include prior cannabis use and perceived efficacy of medical cannabis in relieving PTSD symptoms. Prior cannabis use had a mean accuracy reduction of 0.022 (95% CI: 0.002–0.039), while medical cannabis efficacy exhibited a reduction of 0.017 (95% CI: 0.002–0.032). Similar to the pruned decision tree, the variables sex, race, PTSD severity, and everyday cannabis use, among others, showed no reduction in accuracy when permuted (mean accuracy reduction = 0), indicating that these factors were not critical to the predictive performance of the decision tree in this analysis.

## 4 Discussion

In this study, among the 970 participants surveyed, 62.7% reported being prescribed benzodiazepines prior to obtaining their medical cannabis card, and over half of these individuals (55.8%) = discontinued their benzodiazepine treatment after starting to use medical cannabis. We identified predictors of BZD discontinuation among medical cannabis patients finding that psychiatric care, cannabis use prior to receiving a medical card for accessing medical cannabis, perceived efficacy of cannabis for relieving PTSD symptoms, and traumatic brain injury (TBI) history were critical in predicting discontinuation. The decision tree highlighted that receiving psychiatric care was the most important factor, while the interaction between prior cannabis use and psychiatric care revealed nuanced patterns in the logistic regression model. Together, these results provide insight into the factors that influence BZD discontinuation in a population using medical cannabis for therapeutic purposes.

The pruned decision tree model indicated that not receiving psychiatric care was the strongest predictor of BZD discontinuation, contributing to a notable reduction in model accuracy when permuted. Patients not receiving psychiatric care were more likely to discontinue BZDs, potentially indicating a lower reliance on psychiatric interventions. However, this finding appears to be complex. Takeshima et al. (2022) found that psychiatrists face significant challenges when attempting to discontinue BZDs in patients, particularly due to the recurrence of psychiatric symptoms such as anxiety and insomnia during the tapering process. The study showed that while psychiatrists tend to implement more strategies, such as gradual tapering and psychosocial therapies, to aid in BZD discontinuation, these methods were often complicated by patients’ symptom relapse during the tapering process. Moreover, those currently receiving psychiatric care may not have access to medical cannabis, or are strongly advised not to use medical cannabis by their physicians.

Additionally, those who used cannabis prior to obtaining a medical card emerged as a significant predictor, particularly in combination with psychiatric care as shown by the interaction term. Patients who had used cannabis before obtaining a medical cannabis card were less likely to discontinue BZDs if they were also receiving psychiatric care. This interaction suggests that the therapeutic pathways and expectations related to cannabis use differ based on the type of care patients receive.

Our logistic regression results further underscored the role of psychiatric care and prior cannabis use. The model revealed that prior cannabis use alone was not a significant predictor of discontinuation, but when paired with psychiatric care, the odds of discontinuation dropped substantially. This finding suggests that the effects of cannabis as an alternative therapeutic option may be moderated by the type and intensity of psychiatric interventions patients receive. It also raises important questions about the interplay between traditional psychiatric treatments and emerging therapies such as medical cannabis, which could either complement or compete with conventional treatments depending on the patient’s context.

These results must be placed in the broader context of the health benefits of BZD discontinuation. Long-term use of BZDs is associated with adverse effects (Guina et al., 2015; U.S. Department of Veterans Affairs, 2022), particularly among older adults, including an increased risk of cognitive decline, falls, fractures, and dependence (Takamine et al., 2024). Previous research has highlighted the challenges of discontinuation, noting that patients often experience significant anxiety and withdrawal symptoms during tapering (Maust et al., 2023; Reid Finlayson et al., 2022; Takamine et al., 2024; Takeshima et al., 2022; Fernandes et al., 2022). Participants were not instructed or required to discontinue their BZDs nor was their medical cannabis card approved for the purposes of BSD discontinuation. Rather, they chose to do so after initiating medical cannabis treatment. This decision is especially striking given the difficulties frequently reported in the literature around tapering off these medications (Maust et al., 2023; Reid Finlayson et al., 2022). The decision tree analysis revealed that patients who found their medical cannabis treatment to be highly effective for their PTSD symptoms were more likely to discontinue BZDs, suggesting that medical cannabis may serve as a viable alternative for managing BZD withdrawal symptoms.

Patients who perceived cannabis as effective in relieving their PTSD symptoms were significantly more likely to discontinue BZDs, a finding consistent with previous research on the therapeutic effects of cannabinoids on PTSD (Rehman et al., 2021; Pillai et al., 2022; Nacasch et al., 2023; Krediet et al., 2020; Bonn-Miller et al., 2014; Chan et al., 2017; Drost et al., 2017). Cannabis has been shown to reduce symptoms such as hyperarousal, anxiety, and sleep disturbances, which are often the primary reasons for BZD prescriptions. By offering a safer alternative with fewer long-term risks, medical cannabis may serve as a valuable tool in both managing PTSD and facilitating the discontinuation of more harmful sedative medications.

Balancing the potential benefits of medical cannabis in facilitating BZD discontinuation with the risk of adverse events is crucial for optimizing patient outcomes. While our findings support the use of medical cannabis as an alternative for managing withdrawal symptoms and reducing reliance on BZDs, it is important to recognize that medical cannabis use is not without risks. Adverse events, such as cognitive impairment, dependency, and potential psychiatric effects, must be considered. Clinicians should weigh these potential risks against the known harms of long-term BZD use, especially in vulnerable populations like PTSD patients. A thorough assessment of individual patient profiles, including their history with psychiatric care and their perception of cannabis efficacy, is essential in determining the appropriateness of medical cannabis as part of a broader therapeutic strategy.

Additionally, these findings also raise important clinical questions about the integration of medical cannabis into psychiatric care. The interaction between prior cannabis use and psychiatric care suggests that medical cannabis may not be equally effective for all patients, particularly those who are heavily reliant on traditional psychiatric interventions. Clinicians should carefully assess the therapeutic goals and expectations of patients using medical cannabis, ensuring that it is integrated into a comprehensive care plan that takes into account both the potential benefits and limitations of cannabis as a treatment option.

### 4.1 Limitations

There are several limitations to consider. First, this study relied on self-reported measures, which can introduce various forms of bias (e.g., recall bias). While we did implement follow-up procedures and incentives, our overall response rate was relatively low (10.6%). Analysis of respondent vs. non-respondent demographics showed no significant differences in age or veteran status, but a higher proportion of male and White, non-Hispanic participants responded to the survey. These differences may limit the generalizability of our findings, particularly if gender- or race-based factors shape patients’ decisions regarding benzodiazepine discontinuation or medical cannabis usage.

Additionally, our data do not distinguish between medical vs. recreational cannabis consumption outside of the medical card framework. Consequently, we cannot definitively conclude whether observed benzodiazepine discontinuation was driven predominantly by medical cannabis or other personal or clinical factors. Lastly, patients were not instructed or required to discontinue BZDs, so causal inferences about medical cannabis’s role in prompting discontinuation should be approached with caution. Despite these limitations, this study provides a critical first look at the relationship between medical cannabis use and BZD discontinuation in the PTSD population, underscoring the need for future research using more robust designs such as longitudinal cohorts or electronic health record linkage studies.

### 4.2 Conclusion

This study demonstrates that medical cannabis holds promise as an alternative therapy that can aid in the discontinuation of BZDs, particularly for patients with PTSD. The results suggest that psychiatric care, prior cannabis use, and perceptions of cannabis efficacy are key factors in predicting discontinuation. Given the health risks of long-term BZD use, these findings underscore the importance of exploring alternative treatments like medical cannabis that could provide safer, long-term solutions. Future research should continue to investigate the complex interplay between cannabis use and psychiatric care to optimize treatment strategies for patients seeking to discontinue BZDs.

## Data Availability

Aggregated data is available upon reasonable request to the authors.

