## Supplemental Material for "Identifying Predictors of Benzodiazepine Discontinuation in Medical Cannabis Patients with Post-traumatic Stress Disorder Using a Machine Learning Approach"

**Supplemental Figure 1**: Initial decision tree estimating the most influential predictors of benzodiazepine discontinuation among medical cannabis patients with post-traumatic stress disorder

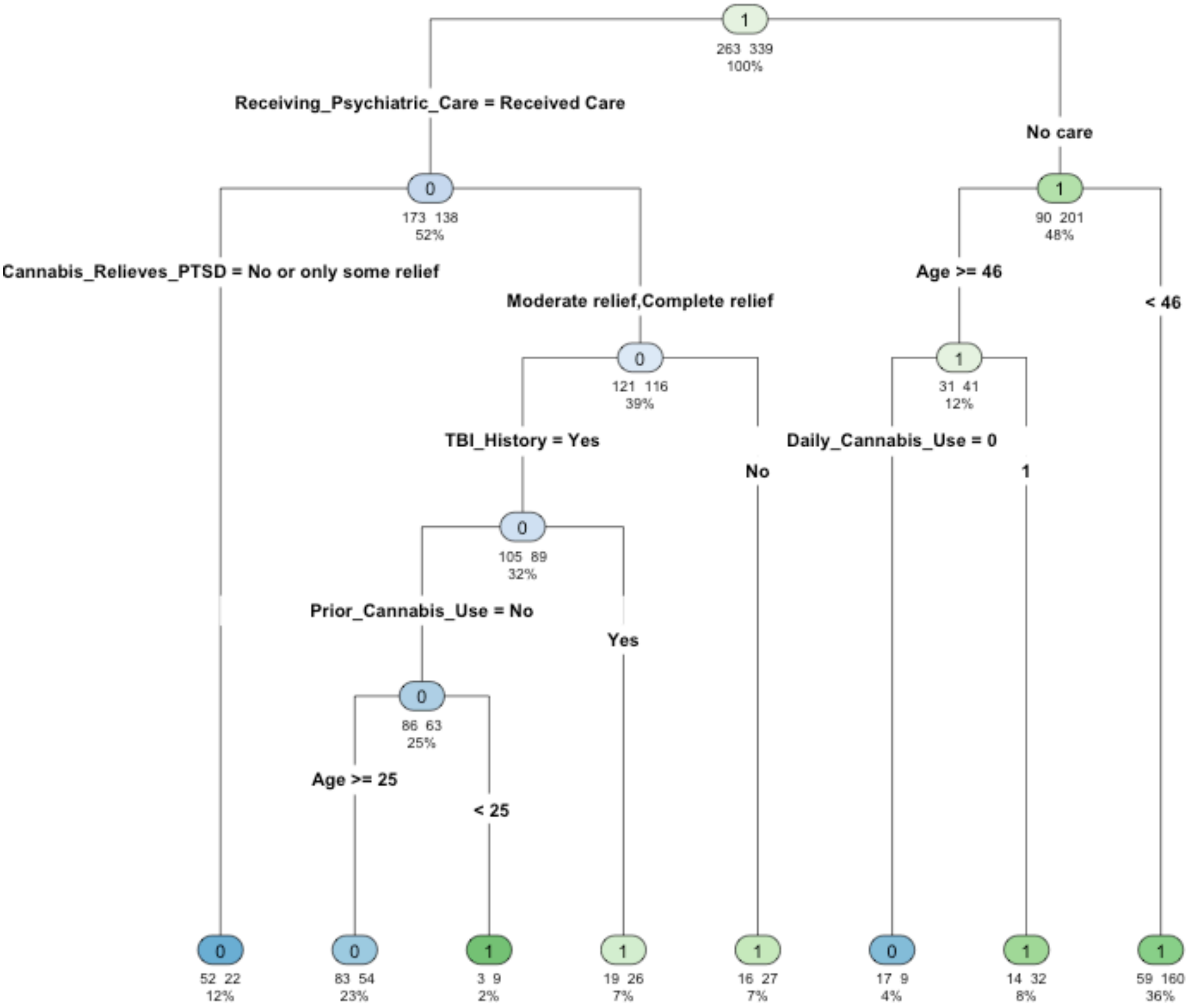

**Supplemental Figure 2**: Visualization of the complexity parameter cross-validation table for the initial decision tree

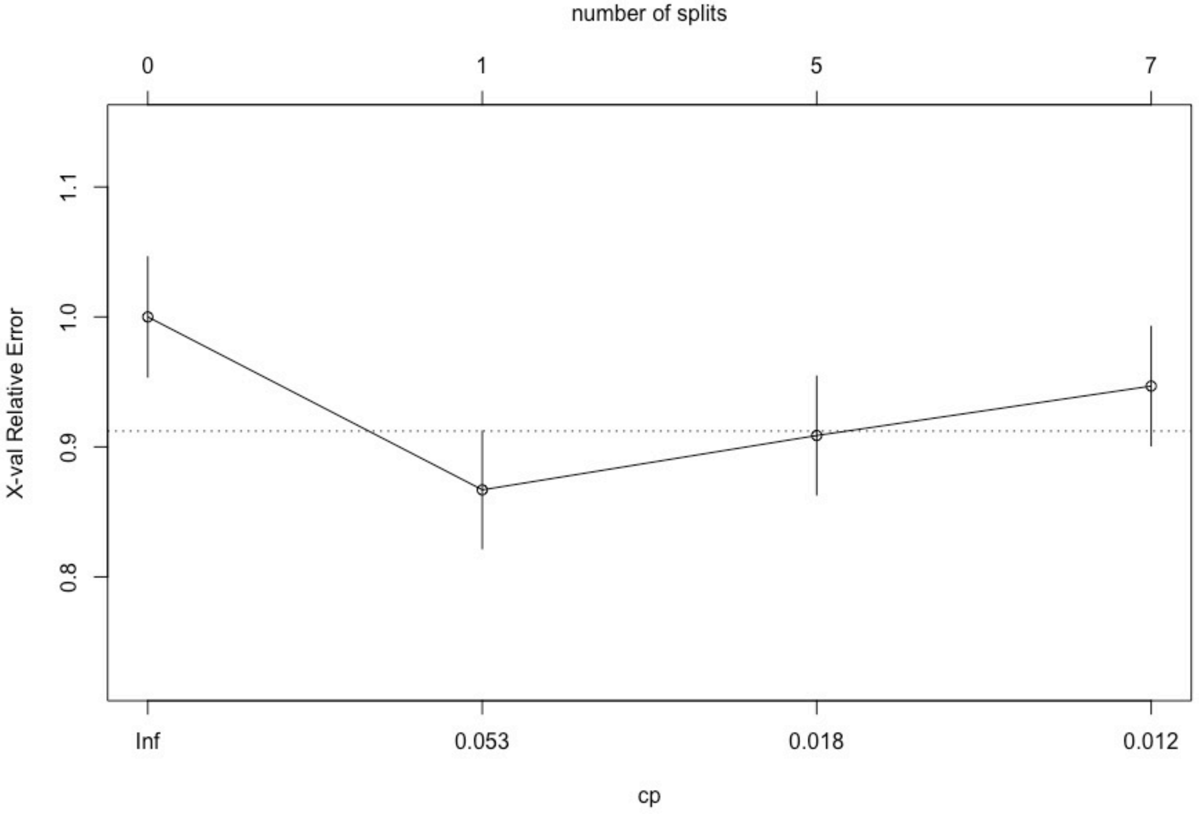

**Supplemental Table 1**: Comparison of respondents versus non-respondents on available demographics

| | | Respondents | Non-respondents | p-value |
|---|---|---|---|---|
| | | n (%)<br><br>*Mean (Standard Error (SE)* | n (%)<br><br>*Mean (SE)* | |
| **N*** | | 970 | 8,198 | |
| **Demographics** | | | | |
| *Gender* | | | | |
| | Female | 305 (31.5%) | 4,442 (54.2%) | <0.001 |
| | Male | 630 (65.1%) | 3,681 (44.9%) | |
| | Other gender listed | 33 (3.4%) | 75 (0.92%) | |
| *Age* | | *40.02 (0.44)* | *39.32 (0.096)* | 0.9323 |
| *Race, Categorical* | | | | |
| | White, non-Hispanic | 695 (73.8%) | 5,364 (68.4%) | 0.001 |
| | Black, non-Hispanic | 96 (10.1%) | 1,138 (14.5%) | |
| | Hispanic | 80 (8.4%) | 643 (8.2%) | |
| | Other Race Listed | 79 (8.3%) | 698 (8.9%) | |
| *Veteran status* | | | | |
| | Yes | 134 (13.8%) | 930 (14.3%) | 0.676 |
| | No | 834 (86.2%) | 5,572 (85.7%) | |

**Note:** *Among respondents, there were 2 missing data points for gender, 20 missing data points for race/ethnicity, and 2 missing data points for veteran status. Among non-respondents, there were 355 missing data points for race/ethnicity and 1,696 missing data points for veteran.

**Supplemental Table 2**: Summary statistics for pruned decision tree estimating the most influential predictors of benzodiazepine discontinuation among medical cannabis patients with post-traumatic stress disorder

| **Statistics** | **Values** |
|---|---|
| Accuracy | 0.6611 |
| Accuracy, Lower 95% Confidence Interval (CI) | 0.6218 |
| Accuracy, Upper 95% CI | 0.6989 |
| No information rate (NIR) | 0.5631 |
| P-value [Accuracy > NIR] | 0.0000 |
| Kappa | 0.2957 |
| McNemar P-value | 0.0004 |
| Sensitivity | 0.5133 |
| Specificity | 0.7758 |
| Positive Predictive Value | 0.6398 |
| Negative Predictive Value | 0.6726 |
| Precision | 0.6398 |
| Recall | 0.5133 |
| F1 | 0.5696 |
| Prevalence | 0.4369 |
| Detection Rate | 0.2243 |
| Detection Prevalence | 0.3505 |
| Balanced Accuracy | 0.6446 |

**Supplemental Table 3**: Permutation test results for variable importance in predicting benzodiazepine discontinuation using a pruned decision tree model

| Variable | Mean Accuracy Reduction | Lower 95% Confidence Interval (CI) | Upper 95% CI |
|---|---|---|---|
| Age | 0.012 | 0.002 | 0.022 |
| Anxiety Severity | 0.000 | 0.000 | 0.000 |
| Relief from PTSD symptoms after using medical cannabis | 0.018 | 0.002 | 0.035 |
| Daily Cannabis Use | 0.000 | 0.000 | 0.000 |
| Education level | 0.000 | 0.000 | 0.000 |
| Experienced Violence in Life? | 0.000 | 0.000 | 0.000 |
| Gender | 0.000 | 0.000 | 0.000 |
| Currently receiving group therapy? | 0.000 | 0.000 | 0.000 |
| Currently receiving individual therapy? | 0.000 | 0.000 | 0.000 |
| Insomnia Severity | 0.000 | 0.000 | 0.000 |
| Living in state with medical cannabis only | 0.000 | 0.000 | 0.000 |
| Cannabis use prior to obtaining a medical card | 0.021 | 0.003 | 0.040 |
| PTSD Severity | 0.000 | 0.000 | 0.000 |
| Race/Ethnicity | 0.000 | 0.000 | 0.000 |
| Currently Receiving Psychiatric care? | 0.120 | 0.086 | 0.151 |
| PTSD Symptom | 0.000 | 0.000 | 0.000 |
| Experience Traumatic Brain Injury? | 0.016 | 0.000 | 0.030 |
| Veteran Status | 0.000 | 0.000 | 0.000 |
